# Upper airway ciliary dysfunction in bronchiectasis: The EMBARC cilia cohort study

**DOI:** 10.64898/2026.02.03.26345362

**Authors:** Mathieu Bottier, Erin Cant, Lidia Perea, Morven K. Shuttleworth, Mahmoud Fassad, Hannah M Mitchison, Stefano Aliberti, Pieter C Goeminne, Holly Lind, Kateryna Viligorska, Emma D Johnson, Jaaming New, Merete B Long, Josje Altenburg, Michal Shteinberg, Francesco Blasi, Oriol Sibila, Eva Polverino, Claire Hogg, Sarah Ollosson, Michael R Loebinger, Natalie Lorent, James D. Chalmers, Amelia Shoemark

## Abstract

Mucociliary clearance is a key component of the pathophysiology of bronchiectasis but cilia function is poorly defined.

This study aims to characterize nasal ciliary function in bronchiectasis and examine associations with disease severity, infection, inflammation and outcome.

Adults with bronchiectasis and healthy volunteers were recruited to the international observational study EMBARC-BRIDGE. Individuals with a known diagnosis of Primary Ciliary Dyskinesia (PCD) were excluded. Nasal respiratory epithelium was sampled by brush biopsy. Ciliary function was assessed by high-speed video microscopy in primary samples and following re-differentiation in air-liquid interface (ALI) culture. Ciliary parameters (cilia length, angle, amplitude, clearance, frequency and ciliation) were quantified and compared with disease severity, microbiology, inflammation and future risk of exacerbations.

171 participants with bronchiectasis were recruited (54% female, age 68years (59-74)). Bronchiectasis nasal brushings showed greater epithelial disruption compared to healthy volunteers (p=0.0006). Six individuals with previously undiagnosed PCD were identified and excluded. In the remaining bronchiectasis cohort, ciliary beat frequency and length were similar to healthy controls. In contrast ciliary beat amplitude, angle, amplitude per second and clearance capacity, were significantly reduced (all p<0.001). These parameters were restored following ALI culture. Regenerated epithelia from bronchiectasis donors exhibited reduced ciliated area. Ciliary dysfunction was strongly associated with future risk of severe exacerbations.

The upper airway epithelium is disrupted in bronchiectasis; ciliary movement is impaired and is associated with future risk of exacerbation. Ciliary dysmotility is reversible following ALI culture. This indicates that impaired ciliary function is secondary to the airway environment and therapeutically targetable.

## Introduction

Bronchiectasis is a heterogeneous disease characterised by chronic cough, sputum production, and exacerbations. The vicious vortex model of disease describes a complex interplay between dysfunction of mucociliary clearance, chronic bacterial infection, and chronic inflammation, which drives and perpetuates disease (1).

Despite the clear implication of mucociliary dysfunction in the pathogenesis of bronchiectasis through inherited diseases such as Primary Ciliary Dyskinesia (PCD) and Cystic Fibrosis (CF), little is known about ciliary function in bronchiectasis of other aetiologies. Improved understanding of infection and inflammatory pathways in bronchiectasis have led to identification of disease endotypes and new avenues for treatments (2-5). Whereas ciliary function, a key potential target for treatment, remains relatively unexplored. To date, only four studies have examined ciliary function in bronchiectasis. The most recent identified a reduction in dynein arm proteins, the motor proteins which drive ciliary beating, in nasal epithelium by immunofluorescence, suggesting direct ciliary defects (6). Functional studies of nasal ciliary beat frequency (CBF) from previous decades have yielded conflicting results (7),(8), (9). The use of high-speed video microscopy and advanced analysis to measure cilia function in depth may overcome limitations of these prior studies and provide tools to measure ciliary function in disease management and the development of new therapies.

30 to 70% of patients with bronchiectasis have upper airway symptoms consistent with chronic rhinosinusitis, and upper airway ciliary function has been shown to correlate with lower airway function (10, 11). Most recently, upper airway microbiome changes were found to correlate with airway infection status and predict future exacerbations (12). Upper airway samples are more accessible than bronchoscopy specimens making them an attractive target to assess cilia function in bronchiectasis patients.

The aim of this study was to investigate the characteristics and reversibility of nasal ciliary dysfunction in adults with bronchiectasis, and to assess how ciliary movement parameters relate to clinical features, airway infection, and inflammatory profiles.

## Methods

### Study design

Patients with bronchiectasis were enrolled in a cilia sub-study of the observational cohort study EMBARC-BRIDGE (NCT03791086). Patients were recruited at Ninewells Hospital, University Hospitals Leuven, Royal Brompton Hospital, and the Hospital de la Santa Creu i Sant Pau between August 2019 and June 2022 (with a 1-year interruption between March 2020 and March 2021 due to the COVID-19 pandemic). The inclusion criteria were: a CT scan showing bronchiectasis along with a compatible clinical syndrome of cough, sputum production and/or recurrent respiratory tract infections and with a primary diagnosis of bronchiectasis made by a respiratory physician; clinically stable state as indicated by the lack of any treatment with antibiotics or corticosteroids for a pulmonary exacerbation in the previous four weeks. Exclusion criteria were inability to give informed consent, <18 years of age, patients with active tuberculosis, people with bronchiectasis due to CF. Exclusion criteria specific to the cilia sub-study comprised: people with a known diagnosis of PCD, active smoking status, an acute upper respiratory tract infection within 6 weeks of the visit (due to known effects on mucociliary function); people with known blood clotting disorders; and individuals currently taking anti-coagulants were excluded for safety.

Participants underwent a nasal brushing for laboratory analysis of cilia function on a single visit. Disease characteristics were recorded using the EMBARC registry as previously described (9). The Bronchiectasis Severity Index (BSI) and FEV1% predicted were measured to evaluate disease severity (10). Sputum neutrophil elastase activity was used as a biomarker of disease activity (11). Eosinophilic inflammation was defined as blood eosinophils >300 cells/µL (12). A history of *Pseudomonas aeruginosa* in sputum and exacerbations in the past year were recorded. Participants were followed up from time of cilia assessment until January 2025 for severe exacerbations during follow-up defined as requirements for hospitalization or intravenous antibiotics. Physician diagnosed chronic rhinosinusitis with and without nasal polyps were also recorded.

Healthy volunteers were required to be non-smokers with no history of respiratory disease. As there was no association with cilia function and age or sex the population was not age or sex matched.

Patients with bronchiectasis contributed to the design of this study through the European Lung Foundation/EMBARC Patient Advisory Group (13).

### Ethics

The protocol was approved by the London - Chelsea Research Ethics Committee (18/LO/1935). All participants provided informed written consent to participate.

### Laboratory analysis

#### High Speed Video-Microscopy (HSVM)

Nasal epithelial cells in suspension in culture media were recorded at a framerate of 500 frames/seconds for 2 seconds. A minimum of 8 side views and 2 top views from independent ciliated epithelial strips were recorded and analyzed.

#### Ciliary function analysis

Videos of ciliated epithelial strips recorded by HSVM were analyzed using an in-house designed Matlab R2022a (The MathWorks, Inc, Natick, MA, USA) program (14, 15). Briefly, position of the cilium base and extreme positions of the cilium tip at the start and end of an active phase of a cycle of beating were recorded. Ciliary Beat Frequency (CBF) was calculated by Fast Fourier Transform and video-kymography. Ciliary beat pattern parameters such as cilium length, angle of beating and beating amplitude were calculated from tip and base positions (Figure 1a). Ciliary beat amplitude per second was calculated as beating amplitude × CBF. Epithelial disruption was scored based on categories described by Thomas (16): 1. normal, 2. minor disruption, 3. major disruption and 4. single cells. Clearance was measured using optical flow by a program designed inhouse and run in Python programming language (Python Software Foundation, version 3.13).

**Figure 1:**
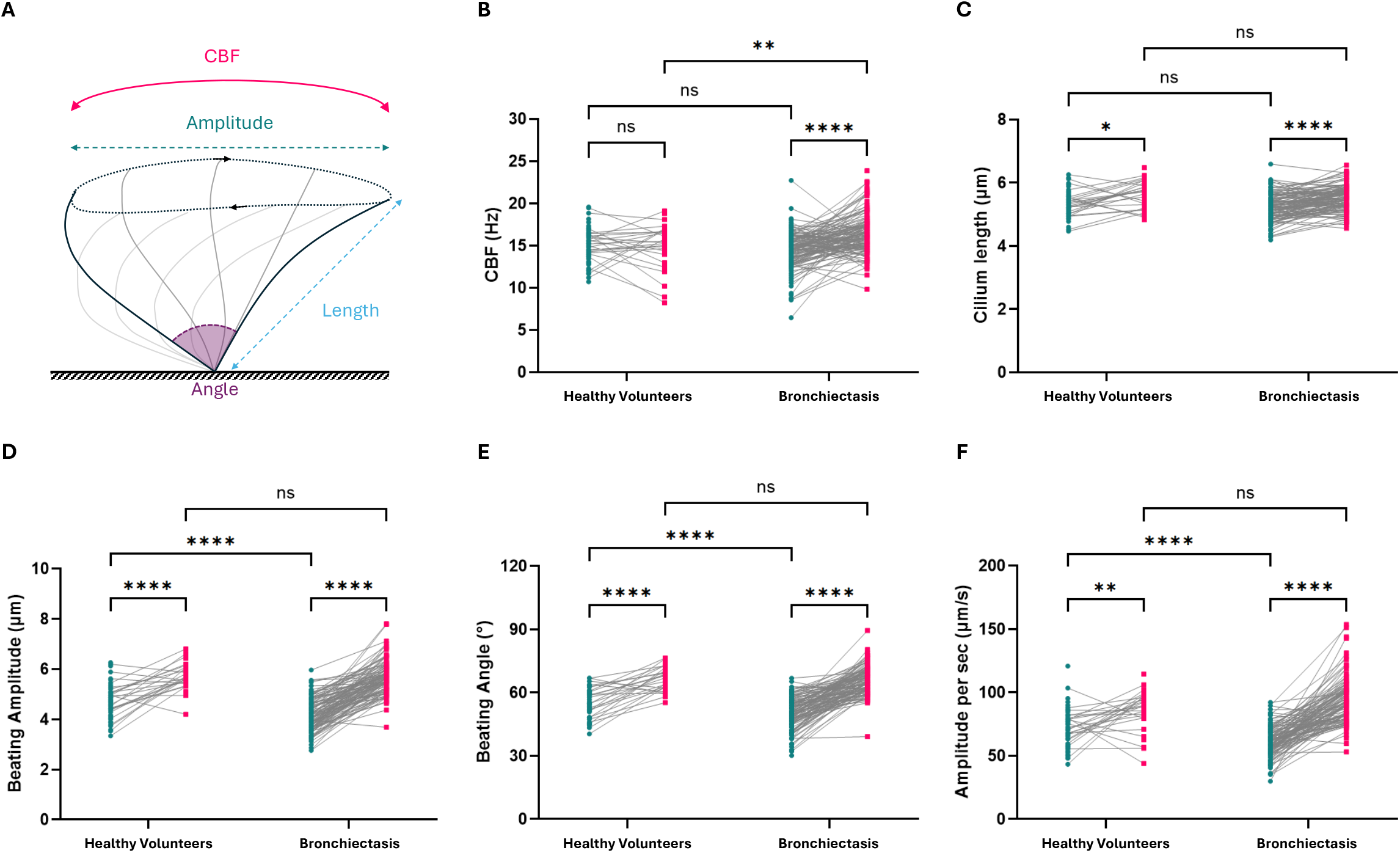
Quantitative analysis of ciliary beating in healthy volunteers and patients with bronchiectasis using high-speed video microscopy. (A) Schematic representation of ciliary motion parameters measured: ciliary beat frequency (CBF), beating amplitude, beating angle, and cilium length. (B–F) Paired analysis of individual subjects before (teal) and after (pink) culturing in control and bronchiectasis groups. (B) Ciliary Beat Frequency (CBF, Hz); (C) Cilium length (μm); (D) Beating amplitude (μm); (E) Beating angle (°) and (F) Beating amplitude per second (μm/s). Grey lines connect paired samples. Paired comparisons using Wilcoxon test and group comparisons using Mann-Whitney (****p < 0.0001, **p < 0.01, *p<0.05, ns = not significant).

#### Air liquid interface culture

Basal cells were isolated from nasal brushings, expanded to passage 2 and differentiated into ciliated epithelium at Air Liquid Interface (ALI). High speed video parameter analysis was repeated at day 37 (+/-5 days) (17).

#### Ciliation quantification

To quantify the level of ciliation post culture, cells were scraped and dried on slides overnight. Slides were stained for immunofluorescence as previously described for acetylated tubulin to visualize cilia and TP63 to visualize basal cells and dapi to mark the cell nucleus. Cell numbers and fluorescence intensity were quantified in Image J (18).

Further detailed methods for cilia analysis methods are available in the supplementary information

#### Data analysis

Cilia function parameters were analyzed using Mann-Whitney or Wilcoxon signed-rank test analysis for group comparisons; Spearman nonparametric correlation for parameters correlation; Fisher’s exact test for population distribution. All tests were carried out with software GraphPad Prism version 9.3.1 for Windows (GraphPad Software, San Diego, CA, USA). For analysis of future severe exacerbations, negative binomial regression was used, with time in follow-up as an offset. Analysis was performed in SPSS version 30 (IBM). For all analyses, a *p*-value < 0.05 was considered statistically significant. Data are shown as mean ± std or median IQR as appropriate.

## Results

### Study population

One hundred and seventy-one subjects with bronchiectasis including 92 female (54%) were enrolled in the EMBARC-BRIDGE cilia study. The age was 68 years (59-74) (median (Q1-Q3). Clinical features are shown in Table 1. The cohort was representative of the wider European EMBARC registry cohort (19). Seventy-three healthy volunteers (53% female) were recruited with average age 54 years (30-69). There was no relationship between age or sex and ciliary function parameters within the healthy controls or overall (supplementary table).

**Table 1.**
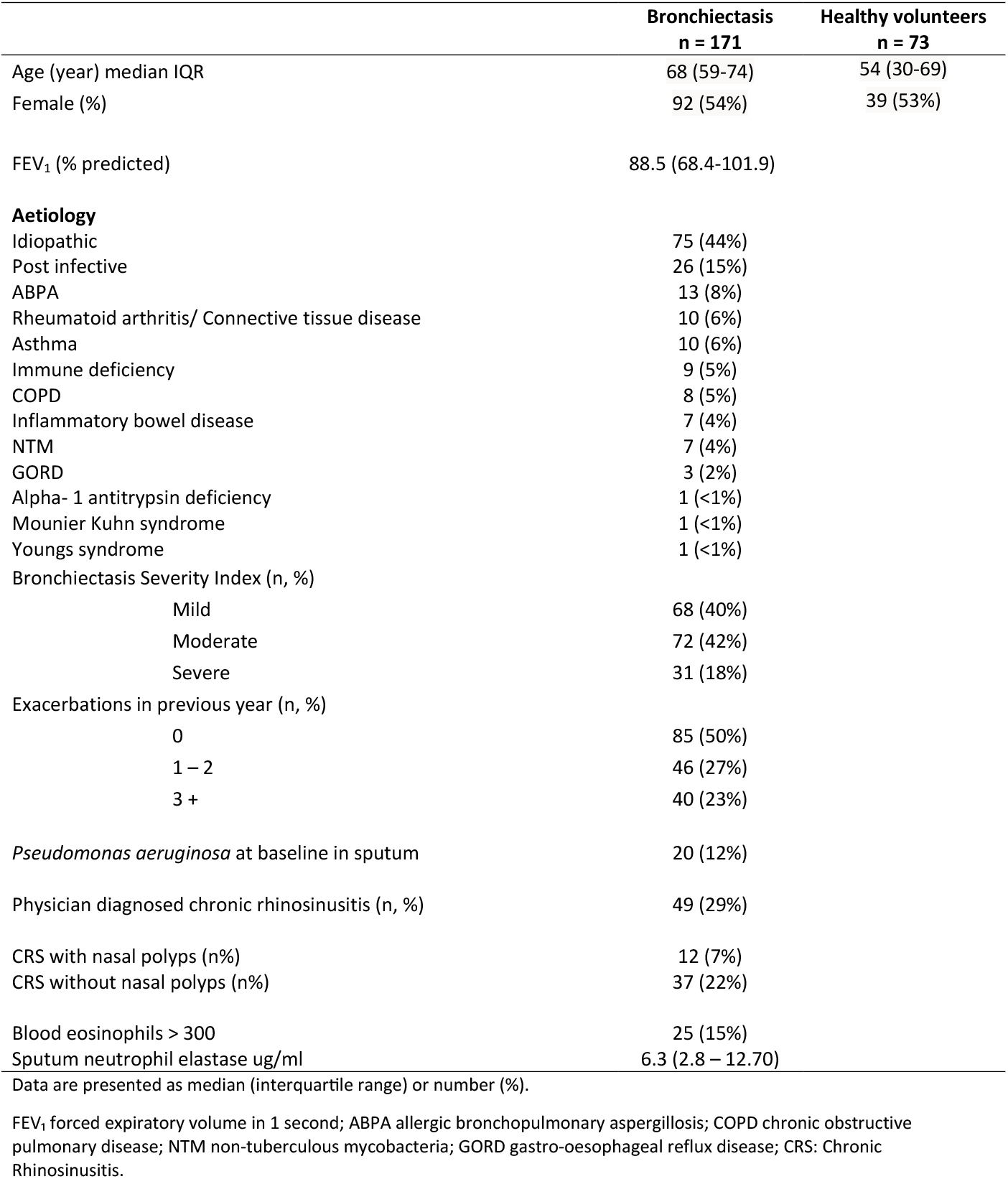
Demographics and Clinical characteristics of patients with bronchiectasis and healthy volunteers.

149/171 bronchiectasis patients (87.1%) had a successful nasal brushing with sufficient epithelial strips for HSVM. 134 samples (78.4% of subjects) were successfully re-differentiated into ciliated epithelium at air liquid interface culture and analyzed.

### Identification of undiagnosed PCD

A known diagnosis of PCD was an exclusion criterion of this study. However, six participants of the 134 with successful ALI culture (4.5%) had a ciliary beat pattern abnormality known to be associated with PCD which persisted following re-growth of the epithelium in vitro. All 6 participants were followed up with whole exome sequencing which revealed a genetic diagnosis in 3 of the 6 (biallelic or male X-chromosome pathogenic variants in *OFD1, RSPH9 and DNAH11*). Gene defect was consistent with known genotype and cilia function phenotype. The remaining 3 were diagnosed with Primary Ciliary Dyskinesia (PCD) as ‘highly likely or suspected PCD’ according to ERS and ATS guideline definition and all 6 were excluded from further analyses (20). Results are shown in Table 2 and Supplementary Videos 1-6.

**Table 2.** Diagnostic findings for 6 participants diagnosed with PCD. Beat pattern abnormality present in both the primary sample and after ALI culture

|  | Beat pattern abnormality | Whole exome sequencing variants | Additional information | ERS/ATS Guideline diagnosis |
| --- | --- | --- | --- | --- |
| **PCD1** | Rotation | Negative for pathogenic or likely pathogenic variants in known PCD genes | Low nasal nitric oxide, class 2 central complex defect by electron microscopy | PCD highly likely |
| **PCD2** | Distal ciliary stasis, abnormal beat pattern | Negative for pathogenic or likely pathogenic variants in known PCD genes | Bronchiectasis in all lobes, pseudomonas, rhinosinusitis and polyps. Electron microscopy normal. | Suspected PCD |
| **PCD3** | Rotation | *RSPH9* (NM_152732)<br>c.804_806del, p.(Lys268del)<br>Homozygous |  | PCD confirmed |
| **PCD4** | Restricted movement of the distal portion of the cilium. 40-60% immotile | *DNAH11* (NM_001277115)<br>c.13456A>T, p.(Arg4486Ter)<br>Homozygous | Electron microscopy showed normal ultrastructure | PCD confirmed |
| **PCD5** | Long cilia with bulbous tips | *OFD1* (NM_003611)<br>c.2745_2746del, p.(Tyr916SerfsTer7)<br>X-linked hemizygous | Electron microscopy revealed sparse cilia and some with long cilia | PCD confirmed |
| **PCD6** | Interrupted beat pattern | Negative for pathogenic or likely pathogenic variants in known PCD genes | Early onset bronchiectasis and a sibling with clinical features of PCD, Normal electron microscopy | Suspected PCD |

### Ciliary function in bronchiectasis compared to healthy volunteers

Bronchiectasis nasal brushings had significantly increased epithelial disruption (friability) scores compared to healthy volunteers (3.0 ± 0.4 vs 2.7 ± 0.6; *p=0*.*0006*), suggesting that the epithelium in people with bronchiectasis is more likely to be friable. This was true of participants both with and without reported Chronic Rhinosinusitis (CRS) (CRS 3.0 ± 0.4; p = 0.0012, No CRS 2.9 ± 0.4; p<0.0001)

We observed clear ciliary dysfunction in the upper airway of bronchiectasis patients compared with healthy volunteers. Comparison of ciliary function between primary nasal brushings of subjects with bronchiectasis and healthy volunteers is shown in Figure 1 and supplementary table 1.

The cilium beating angle, amplitude and amplitude per second and ciliary clearance were significantly reduced in bronchiectasis compared to healthy volunteers (Angle 49.8 ± 9.8 º vs 54.2 ± 11.7 º; Amplitude 4.1 ± 0.9 µm vs 4.6 ± 0.8 µm; Amplitude per second 58.8 ± 14.3 µm/s vs 68.2 ± 19.8 µm/s; Clearance 0.19 ± 0.14 µm/s vs 0.25 ± 0.15 µm/s; *p*<0.0001 all comparisons). Again, there was no difference between patients with and without reported CRS.

Neither ciliary beat frequency nor ciliary length were significantly different between the bronchiectasis and the healthy volunteers group (CBF 14.5 ± 2.6 Hz vs 15.2 ± 2.4 Hz, p = 0.10; Length 5.2 ± 0.5 µm vs 5.3 ± 0.5 µm, p = 0.24) in primary samples.

### Recovery of ciliary function but not epithelial ciliation following in vitro culture at air liquid interface

To assess the impact of the airway environment on ciliary function, cells from 128 patients’ bronchiectasis patients (excluding patients newly diagnosed with PCD) were re-differentiated at air liquid interface (ALI) culture in a sterile environment for over 35 days. The cilium beating angle, amplitude, amplitude per second and clearance parameters significantly increased following ALI culture compared to the paired primary samples (Angle 66.0 ± 9.1 º vs 66.0 ± 9.1 º; Amplitude 5.6 ± 0.8 µm vs 5.7 ± 0.8 µm; Amplitude per second 92.5 ± 20.5 µm/s vs 89.4.± 16.0 µm/s; Clearance 0.19 ± 0.14 µm/s vs 0.37 ± 0.19 µm/s; *p*<0.0001 all comparisons).

After ALI culture, the cilia angle, amplitude, amplitude per second and clearance remained heterogenous but on average were similar between the bronchiectasis and the healthy volunteers groups (p>0.05). (Figure 1D and Supplementary Table 1).

In addition to cilia function, mucociliary clearance is also impacted by ciliary number. The intensity of acetylated α-tubulin fluorescence was used to quantify the level of ciliation after ALI culture. Bronchiectasis cultured epithelium had a significantly lower intensity (53.1 ± 30.3 RFU, n = 80) compared to healthy volunteers cultures (104.7 ± 24.3 RFU, n = 10; *p < 0*.*0001*). Suggesting that despite restoration of cilia function there was reduced ciliation at ALI.

### Nasal ciliary function, disease severity and inflammation

Since epithelial samples were obtained from the upper airway, we assessed upper airway disease defined by symptoms of chronic rhinosinusitis with or without nasal polyps. Chronic Rhinosinusitis was not associated with epithelial disruption, ciliary beat pattern, or mucociliary clearance parameters.

There was no association between ciliary function analysis parameters and disease severity, the latter measured using the Bronchiectasis Severity Index (BSI), or lung function as measured by FEV1 % predicted. HSVA parameters were also not related to prior exacerbation frequency (0, 1–2, or ≥3 exacerbations per year). No relationship was observed with markers of disease activity or inflammation, including sputum neutrophil elastase activity or systemic eosinophilic inflammation as measured by blood eosinophil count. These data are presented in Figure 2 and Supplementary Table 2.

**Figure 2:**
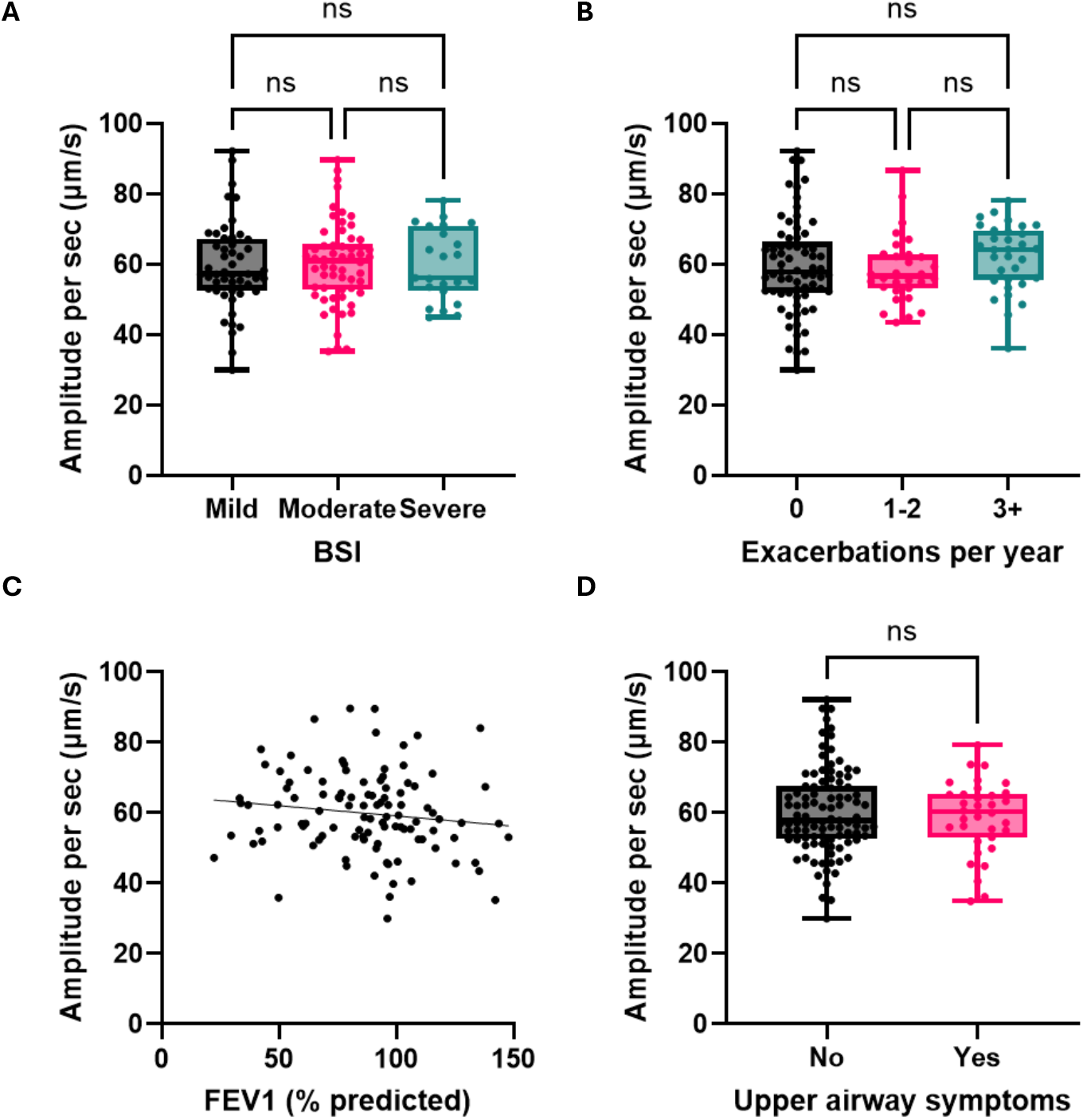
Ciliary beating amplitude per second in bronchiectasis compared to disease severity and activity. (A) Disease severity measured by Bronchiectasis Severity Index (BSI); (B) Lung function by FEV1 (% predicted). Black line shows linear regression (R^2^ = 0.018); (C) No. of exacerbations in the past 12 months; (D) Upper airway disease (CRS with or without nasal polyps); Group comparisons using Kruskal-Wallis and Mann-Whitney (ns = not significant).

While *in vitro* exposure to *Pseudomonas aeruginosa* has been shown to impair ciliary function, no differences in HSVA parameters were observed between individuals who grew *P. aeruginosa* in sputum compared with those who had not (Supplementary Table 2.)

### Nasal ciliary function and future severe exacerbations

Bronchiectasis patients were followed up for between 1 and 3 years for severe exacerbations. 37 severe exacerbations occurred during 354 years of cumulative follow-up. Upper airway ciliary function analysis parameters were clearly associated with future risk of severe exacerbations (Table 3). In primary brushings, findings of longer cilia, reduced beating angle, reduced beating amplitude and amplitude per second, and increased disruption were all associated with an increased frequency of severe exacerbations. Ciliary beat frequency was not associated with severe exacerbations. All data are shown in table 3. No cilia parameters post-culture were associated with severe exacerbations (all *p*>0.05, data not shown) but the 50% of participants with the largest change in amplitude per second had a rate ratio of 4.01 (95%CI 1.36-11.87, *p* = 0.012) compared to patients with a lesser improvement post culture.

**Table 3.**
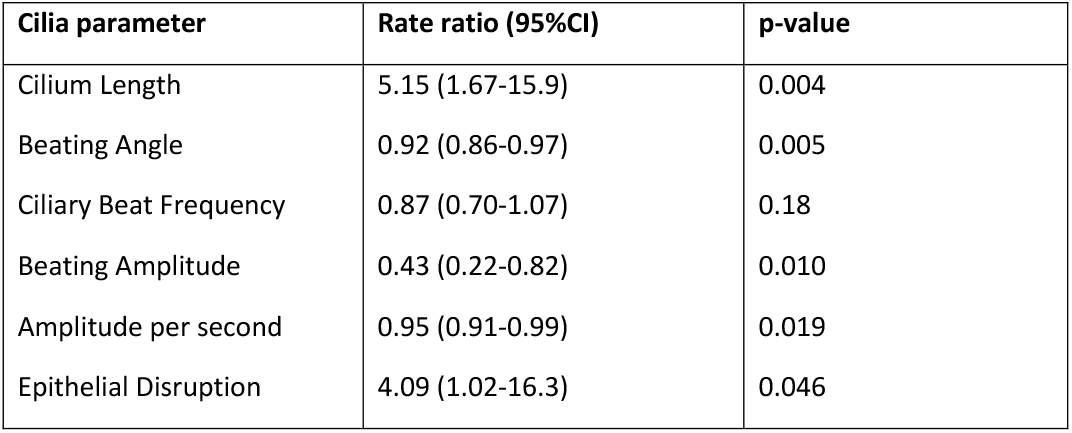
Relationship between ciliary function analysis parameters in primary brushings and future severe exacerbations during follow-up.

## Discussion

This study demonstrates that upper airway integrity, ciliary function and clearance are impaired in people with bronchiectasis, as assessed by high-speed video-microscopy. Abnormalities are not related to physician/patient reported rhinosinusitis and are present across the spectrum of disease severity but are associated with future risk of severe exacerbations. Abnormalities were observed in ciliary beat angle, amplitude, and clearance, while ciliary beat frequency and ciliary length were preserved compared with healthy volunteers. Importantly, these abnormalities were independent of disease severity, infection status, or markers of inflammation, yet predicted future risk. This suggests that ciliary dysfunction is a fundamental feature of bronchiectasis and a marker of the activity of the disease.

A clinically important observation was the identification of six previously undiagnosed cases of primary ciliary dyskinesia (PCD), representing approximately 4.5% of the studied cohort. When combined with individuals with a pre-existing PCD diagnosis within EMBARC who were excluded from this analysis, the estimated prevalence is 7–8%, consistent with prior systematic screening studies in bronchiectasis populations (21, 22). Notably, only half of the newly identified cases had a confirmatory genetic diagnosis. This is not surprising as approx. 30% of cases cannot be solved by current genetics {Legebeke, 2024 #653}. These cases were identified in highly specialized centers and none had classical PCD features such as dextrocardia, in keeping with identification of rarer genotypes. The identification of many PCD patients in adult specialist centers supports the need for wider PCD screening. Underdiagnosis is likely to be even greater outside of specialist centers such as those included in this study. Our findings reinforce the current limitations of genetics as a standalone diagnostic tool for PCD and support the continued need for a multimodal diagnostic approach when PCD is suspected in patients with bronchiectasis (20).

In participants without PCD, ciliary dysfunction was largely reversible following redifferentiation of epithelial cells under sterile air–liquid interface conditions. Restoration of ciliary beat pattern and clearance parameters after regrowth externally from the airway environment strongly suggests that impaired ciliary function in bronchiectasis is predominantly secondary to extrinsic factors rather than intrinsic defects of the cilium. These findings support the concept that the bronchiectasis airway milieu actively suppresses ciliary performance and that this suppression is potentially modifiable, making ciliary function a treatable trait. Several previous studies have shown that infectious and inflammatory stimulus can impact ciliary function (23-26). The recovery of ciliary function observed in vitro is consistent with clinical trial data demonstrating reductions in mucus plugging on high-resolution CT following anti-inflammatory interventions, such as DPP1 inhibition. Although ciliary function normalized, the proportion of ciliated cells remained lower in bronchiectasis cultures than in volunteers, in keeping with studies showing reduced levels of key ciliary proteins by immunofluorescence in primary brushings. However, this is in contrast to recent transcriptomic data demonstrating an upregulated expression of genes involved in ciliogenesis (cilia assembly and organization) in bronchial brushings from patients with radiological, but not clinical bronchiectasis (6, 27). This could be related to differences between methodologies, sampling site and presence of active disease across studies. Larger proteomic and transcriptomic studies of the airway epithelium are needed to confirm these findings.

In contrast to much of the existing literature, which has focused predominantly on ciliary beat frequency, this study highlights the importance of more detailed assessment of ciliary motion (7-9). Rapid but dyskinetic beating may be insufficient to generate effective mucus transport, and effective mucociliary clearance depends on coordinated ciliary motion with sufficient stroke amplitude. In the present study measures of ciliary beat amplitude and clearance were more sensitive indicators of dysfunction than frequency alone. This is reflected in our analysis of future severe exacerbations. While limited by a small number of events, we show a clear relationship between cilia parameters and future severe exacerbations. Our data cannot and should not suggest that this is causal. Rather it may suggest that activity of epithelial disease, reflected by greater disruption of the epithelium and reduced ciliary clearance measured by reduced amplitude per second, is linked to future exacerbation risk.

The strengths of this study include its embedded design within the EMBARC registry, allowing for standardized data collection and the use of quantitative, high-resolution methods to assess ciliary function. Several limitations should be acknowledged. It remains uncertain to what extent nasal ciliary function reflects mucociliary clearance in the lower airways, and while nasal sampling is practical and accessible, it may not fully capture lower airway pathology in bronchiectasis. Future studies will include bronchoscopic sampling. In addition, cilia parameter analyses were conducted on cells in suspension and in the absence of a mucus layer and air exposure, meaning that the contribution of the dehydrated and viscous mucus layer to mucociliary dysfunction in bronchiectasis was not directly assessed (28). These factors potentially confound any ciliary motion defects (29). Upper airway disease was physician diagnosed and not objectively measured, future studies could use the SNOT22 or similar, to reduce underreporting of milder nasal symptoms. Finally, as the study was predominantly performed shortly after the COVID-19 pandemic which disrupted recruitment, this may have led to a lower-than-expected number of exacerbations and may limit generalizability.

In summary, this study shows that ciliary dysfunction in the nasal epithelium is a common feature of bronchiectasis, but one that is largely reversible and independent of disease severity. These findings support the concept of mucociliary dysfunction as a potentially modifiable component of bronchiectasis pathophysiology and identify ciliary function by ciliary function analysis as a plausible and measurable outcome for therapeutic strategies aimed at restoring epithelial health and improving mucociliary clearance.

## Data Availability

All data produced in the present study are available upon reasonable request to the authors.

