## Supplementary information for "Upper airway ciliary dysfunction in bronchiectasis: The EMBARC cilia cohort study"

**Supplementary Table 1: Comparison of HSVM parameters in healthy volunteers vs bronchiectasis** (PCD excluded n=6).

| | | Healthy Volunteers | | Bronchiectasis | | Mann-Whitney |
|---|---|---|---|---|---|---|
| | | Median | iQ | Median | iQ | *p-value* |
| Primary | Sufficient Samples (n) | 65 | | 143 | | |
| | Videos per patient (n) | 12.0 | 5.0 | 15.0 | 5.0 | |
| | Epithelial Disruption | 2.7 | 0.6 | 3.0 | 0.4 | ***<0.0001*** |
| | Length (µm) | 5.3 | 0.5 | 5.2 | 0.5 | *0.2443* |
| | CBF (Hz) | 15.2 | 2.4 | 14.5 | 2.6 | *0.0988* |
| | Angle (degrees) | 54.2 | 11.7 | 49.8 | 9.8 | ***<0.0001*** |
| | Amplitude (µm) | 4.6 | 0.8 | 4.1 | 0.9 | ***<0.0001*** |
| | Amplitude per second (um/s) | 68.2 | 19.8 | 58.8 | 14.3 | ***<0.0001*** |
| | Clearance (optical flow) (µm/s) | 0.25 | 0.15 | 0.20 | 0.14 | ***<0.0001*** |
| Culture | Sufficient Samples (n) | 33 | | 128 | | |
| | Videos per patient (n) | 15.0 | 4.0 | 16.0 | 4.0 | |
| | Length (µm) | 5.7 | 0.7 | 5.5 | 0.5 | *0.2402* |
| | CBF (Hz) | 15.3 | 2.6 | 16.2 | 2.6 | ***0.0066*** |
| | Angle (degrees) | 68.0 | 8.9 | 66.0 | 9.1 | *0.6381* |
| | Amplitude (µm) | 5.7 | 0.7 | 5.6 | 0.8 | *0.4979* |
| | Amplitude per second (µm/s) | 89.4 | 16.0 | 92.5 | 20.5 | *0.0774* |
| | Clearance (optical flow) (µm/s) | | | 0.37 | 0.19 | *N/A* |

**Supplementary Table 2: HSVM parameters compared with age and gender in healthy volunteers.**

| **Age** | | | |
|---|---|---|---|
| | **r value** | | **p-value** |
| Amplitude/s (µm/s) | -0.06 | | 0.61 |
| CBF (Hz) | -0.14 | | 0.23 |
| Angle (degrees) | -0.10 | | 0.41 |
| Cilia length (µm) | -0.02 | | 0.89 |
| Clearance (µm/s) | 0.01 | | 0.88 |
| **Sex** | | | |
| | **Male** | **Female** | |
| Amplitude/s (µm/s) | 67.18 (14.5) | 72.81 (13.9) | 0.11 |
| CBF (Hz) | 14.53 (1.89) | 15.38 (2.07) | 0.09 |
| Angle (degrees) | 53.38 (7.07) | 55.55 (6.25) | 0.19 |
| Cilia length (µm) | 5.33 (0.42) | 5.27 (0.39) | 0.54 |

**Supplementary Table 3: HSVM parameters in bronchiectasis compared with disease severity and activity.**

| **Bronchiectasis Severity Index** | | | | |
|---|---|---|---|---|
| | **Mild (0-4)** | **Moderate (5-8)** | **Severe (>9)** | **P value** |
| Amplitude/s (µm/s) | 58.33 (14.79) | 60.23 (11.86) | 59.90 (10.22) | 0.73 |
| CBF (Hz) | 14.20 (3.17) | 14.54 (1.90) | 13.88 (2.19) | 0.51 |
| Angle (degrees) | 48.47 (10.42) | 50.08 (7.39) | 49.53 (6.95) | 0.62 |
| Cilia length (µm) | 5.06 (0.72) | 5.25 (0.46) | 5.33 (0.40) | 0.08 |
| Clearance (µm/s) | 0.20 (0.13) | 0.22 (0.16) | 0.21(0.14) | 0.62 |
| **Annual exacerbations** | | | | |
| | **0** | **1-2** | **3+** | |
| Amplitude/s (µm/s) | 59.76 (14.27) | 60.29 (10.43) | 56.89 (10.75) | 0.52 |
| CBF (Hz) | 14.23 (1.73) | 14.63 (2.36) | 13.76 (2.24) | 0.31 |
| Angle (degrees) | 49.11 (9.28) | 50.32 (6.65) | 48.05 (6.40) | 0.55 |
| Cilia length (µm) | 5.28 (0.47) | 5.20 (0.39) | 5.26 (0.44) | 0.79 |
| Clearance (µm/s) | 0.20 (0.15) | 0.23 (0.15) | 0.23 (0.19) | 0.24 |
| **FEV1% predicted** | | | | |
| | **≥70% pred** | **<70% pred** | | |
| Amplitude/s (µm/s) | 59.9 (13.6) | 58.1 (14.3) | | 0.47 |
| CBF (Hz) | 14.30 (2.65) | 14.05 (2.80) | | 0.31 |
| Angle (degrees) | 49.50 (9.41) | 48.61 (10.20) | | 0.44 |
| Cilia length (µm) | 5.18 (0.69) | 5.14 (0.76) | | 0.66 |
| Clearance (µm/s) | 0.22 (0.13) | 0.21 (0.15) | | 0.56 |
| ***Pseudomonas aeruginosa* at baseline in sputum** | | | | |
| | **No** | **Yes** | | |
| Amplitude/s (µm/s) | 59.57 (11.17) | 56.88 (12.89) | | 0.49 |
| CBF (Hz) | 14.60 (1.92) | 14.15 (1.73) | | 0.40 |
| Angle (degrees) | 49.89 (7.94) | 47.60 (8.03) | | 0.35 |
| Cilia length (µm) | 5.16 (0.43) | 5.34 (0.38) | | 0.14 |
| Clearance (µm/s) | 0.22 (0.14) | 0.22 (0.15) | | 0.85 |
| **Chronic rhinosinusitis (with or without nasal polyps)** | | | | |
| | **No** | **Yes** | | |
| Amplitude/s (µm/s) | 60.26 (12.20) | 58.82 (10.40) | | 0.51 |
| CBF (Hz) | 14.27 (2.32) | 14.72 (1.93) | | 0.25 |
| Angle (degrees) | 49.72 (7.93) | 49.80 (6.01) | | 0.95 |
| Cilia length (µm) | 5.24 (0.46) | 5.18 (0.30) | | 0.41 |
| Clearance (µm/s) | 0.22 (0.16) | 0.17 (0.09) | | 0.47 |
| **Blood eosinophils** | | | | |
| | **<300** | **≥300** | | |
| Amplitude/s (µm/s) | 58.81 (14.53) | 59.81 (9.51) | | 0.67 |
| CBF (Hz) | 14.02 (2.75) | 15.01 (2.56) | | 0.09 |
| Angle (degrees) | 48.92 (9.94) | 49.34 (7.78) | | 0.82 |
| Cilia length (µm) | 5.13 (0.74) | 5.29 (0.39) | | 0.13 |
| Clearance (µm/s) | 0.21 (0.15) | 0.21 (0.11) | | 0.36 |
| **Neutrophil elastase in sputum** | | | | |
| | **Low <20 ug/ml** | **High >20 ug/ml** | | |
| Amplitude/s (µm/s) | 59.50 (14.0) | 57.60 (12.90) | | 0.53 |
| CBF (Hz) | 14.23 (2.96) | 14.08 (2.00) | | 0.24 |
| Angle (degrees) | 49.11 (10.10) | 48.71 (8.10) | | 0.57 |
| Cilia length (µm) | 5.13 (0.78) | 5.22 (0.36) | | 0.71 |
| Clearance (µm/s) | 0.20 (0.15) | 0.19 (0.12) | | 0.84 |

### Supplementary methods

#### Air liquid interface culture

Nasal cells were seeded on a Nunc™ Cell-Culture Treated 6 well plate (Fisher Scientific - UK Ltd., Loughborough, England) treated with Bovine collagen type 1 6mg/ml (Stemcell Technologies UK Ltd., Cambridge, UK) until confluence (6.6 ± 3.1 days). Cells were then transferred onto Costar® 6.5 mm Transwell® with 0.4 µm Pore Polyester Membrane Insert (Corning Life Sciences, Corning, NY, USA) with PneumaCult™-Ex Plus Medium (Stemcell Technologies UK Ltd., Cambridge, UK) both apically and basally until confluence (3.6 ± 2.1 days). Media was removed from the apical surface and PneumaCult™-ALI Medium (Stemcell Technologies UK Ltd., Cambridge, UK) was added basally during differentiation (36.8 ± 5.8 days) and changed 3 times per week. The apical surface of the cells was gently flushed weekly with PBS to remove mucus. Both PneumaCult™-Ex Plus Medium and PneumaCult™-ALI Medium contained Hydrocortisone (Stemcell Technologies UK Ltd., Cambridge, UK), Penicillin- streptomycin (Fisher Scientific) and Primocin® (InvivoGen Europe, Toulouse, France).

#### High-speed video-microscopy

Nasal epithelial cells were placed in Ibidi µ-Slide 8-well chambers (Ibidi GmbH, Gräfelfing, Germany) and imaged using an inverted Leica SP5 microscope (Leica Microsystems Ltd., Milton Keynes, UK) equipped with a 63× oil-immersion objective. High-speed video recordings were acquired using a Promon U1000 camera (AOS Technologies AG, Baden, Switzerland) at 500 frames per second for 2 seconds (1,000 frames total). The resulting pixel size was 0.075 × 0.075 µm. All recordings were performed at 37°C using the microscope environmental chamber, in accordance with published recommendations (Jackson and Bottier 2022).
For each subject, at least 10 independent ciliated epithelial strips were recorded, including a minimum of eight side-view recordings and two top-view recordings.

#### High-speed video-microscopy analysis

High-speed video microscopy (HSVM) recordings were analyzed using an in-house–developed MATLAB R2022a (MathWorks, Natick, MA, USA) program based on previously published methods (1, 2). After user selection, videos were displayed in a dedicated graphical user interface. The user defined a region of interest (ROI) containing beating cilia and drew a line across the ciliary field.

The position of the cilium base and the extreme positions of the cilium tip at the beginning and end of the active phase of a beating cycle were identified by user mouse clicks using dedicated interface controls (Sup Fig. 1A). Ciliary beat frequency (CBF) was calculated by Fast Fourier Transform (FFT) analysis of the ROI, along the defined cilia line and in the line between Base and Extreem1 position. The cilia line was used to generate a video-kymograph by projecting intensity values along the line over time. Kymograph was displayed in a dedicated interface, allowing the user to manually select the number of beating cycles present (Sup Fig. 1B) and automatically calculate CBF. All four CBF measurements are reported. A median CBF is also provided, defined as the median of the four measurements, with the kymograph-derived CBF weighted by being included twice in the median calculation, as this measurement is least affected by background noise.

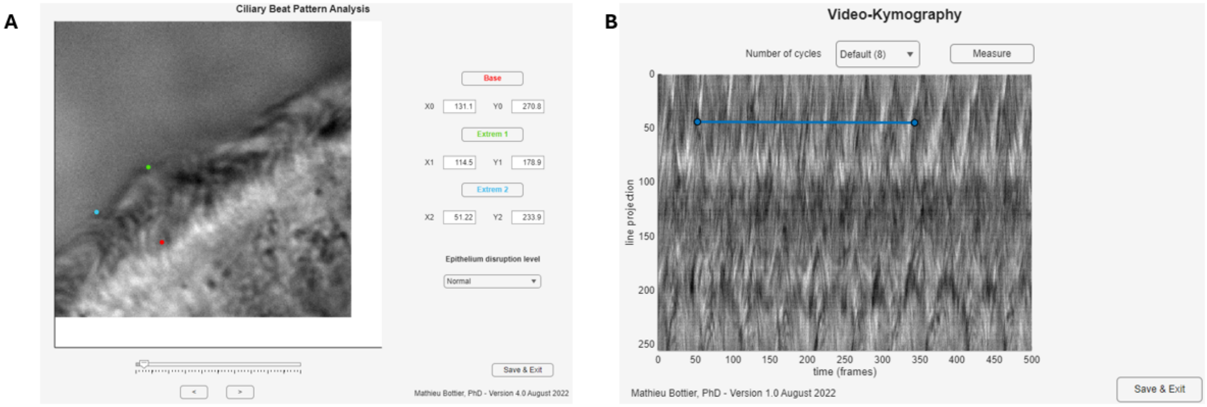


**Supplementary Figure 1 – High Speed Video-Microscopy Analysis.**
(A) User Interface for the analysis of ciliary beating including 3 buttons used to define the position of the base (red) and extreme positions of the cilium tip at the start (green) and end (blue) of an active phase of a cycle of beating. Epithelium disruption level can be selected in the dropdown menu. (B) Video-kymography of the line of pixels selected in the ciliary field and projected over time. The user can select the number of cycle visible and measure the distance (in frames).

Ciliary beat pattern parameters including cilium length, beating angle, and beating amplitude, were calculated using the three user-defined points using basic trigonometric relationships and simple geometric theorems (see Figure 1A in main manuscript). The ciliary beat amplitude per second (defined as beating amplitude × CBF) was used to indirectly describe the muco-ciliary clearance.

Epithelial disruption was scored according to the classification described by Thomas *et al.* (3): (1) normal epithelium defined by an intact and smooth epithelium surface; (2) minor disruption defined by cells projecting out slightly but below the cilia line; (3) major disruption defined by cells projecting out above cilia line; (4) isolated single cells (Sup Fig. 2). Scores were entered via a dedicated drop-down menu within the user interface.

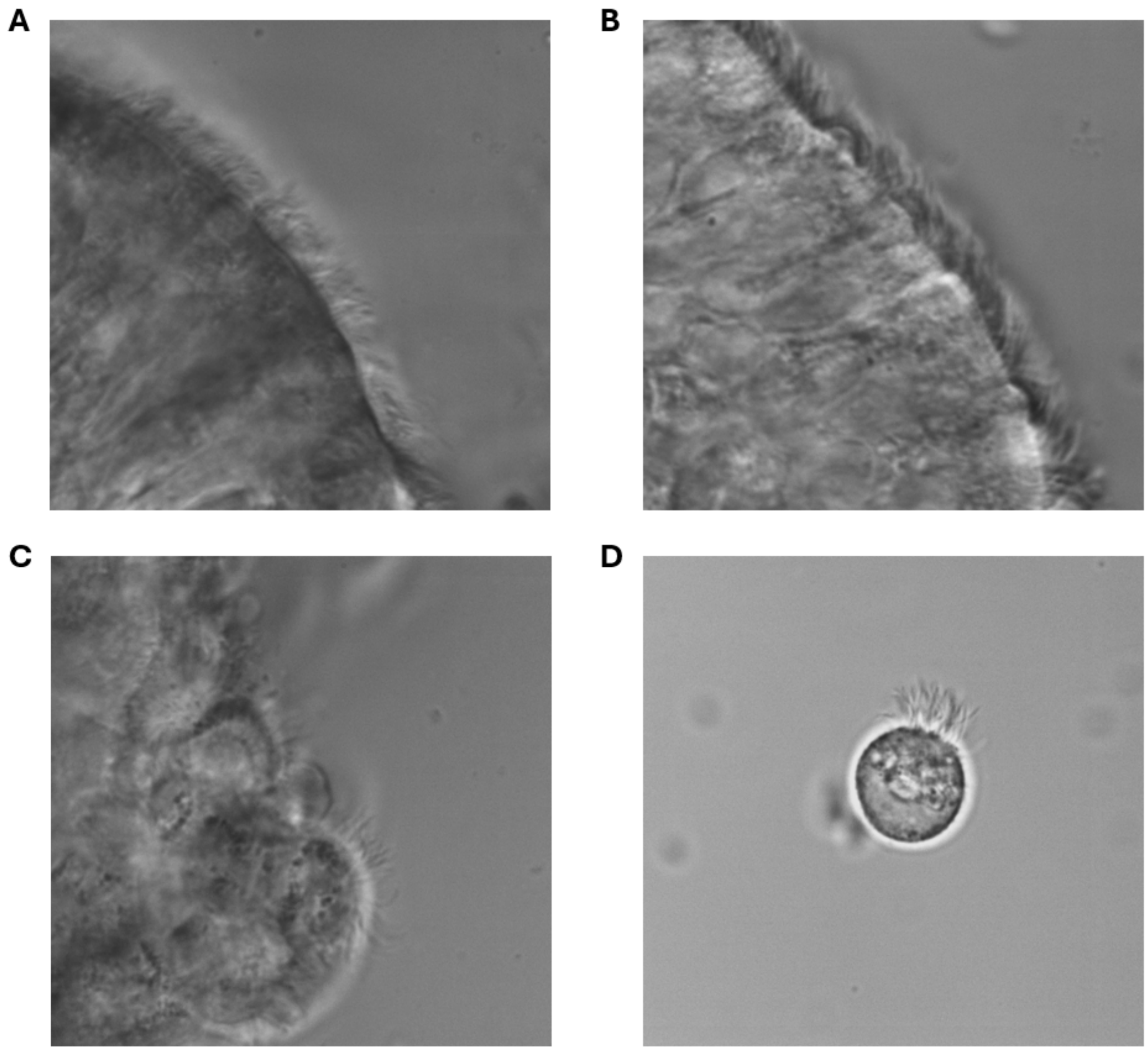


**Supplementary Figure 2 – Examples images of the epithelial disruption score.**
(A) Normal epithelium; (B) Minor disruption; (C) Major disruption; (D) Single cell.

All measured parameters were automatically saved to a comma-separated values (.csv) file. In addition, the corresponding video-kymograph and a figure highlighting the positions of the three selected points were saved in the same directory as the original video files.

#### Optical flow clearance assessment

We used a Python-based method of clearance assessment using optical flow for the quantification of ciliary clearance. The program performs automated analysis of HSVM recordings of ciliated epithelial edges to estimate clearance velocity, using optical flow to measure pixel-level motion across sequential frames converting this motion into a transport speed expressed in micrometres per second (μm/s). The workflow comprised seven sequential steps (Sup Fig 2):

**1. Video Input and Batch Processing** The program automatically scans a selected folder (including subfolders) and processes all .avi video files.

**2. Automatic ROI selection** The program identifies the region of interest (ROI) corresponding to the clearance layer via analysing vertical distribution of motion magnitude. ROI is detected automatically with a confidence score if motion is well defined. If the confidence is low (e.g. noise or insufficient motion), a user prompt asks the observer to check and redefine the ROI or exclude the video from analysis, if for example there are no cilia.

**3. Motion Field Extraction (optical flow)** Using Farneback dense optical flow, the software computes horizontal and vertical displacement vectors between consecutive frames. These displacement fields are averaged across up to the first 500 frames of each video (1 second at a framerate of 500 fps).

**4. Velocity Computation** Motion vectors inside the ROI are converted from pixel displacement to physical transport speed using the known frame rate and pixel-to-micron scaling (0.075 µm/pixel).

**5. Filtering** Videos are filtered for high velocity oscillations (cilia beating) and whole field drift/displacement.

**6. Quality Control Visualizations** To support verification and reproducibility, the software generates diagnostic images per video including a heatmap with ROI boundaries overlaid and a vector field plot showing only transport-aligned motion within the ROI, allowing rapid visual conformation of analysis integrity.

**7. Quantitative Output** For each video, Mean filtered transport velocity (µm/s), ROI detection method and confidence are reported ina comma-separated values (.csv) file.

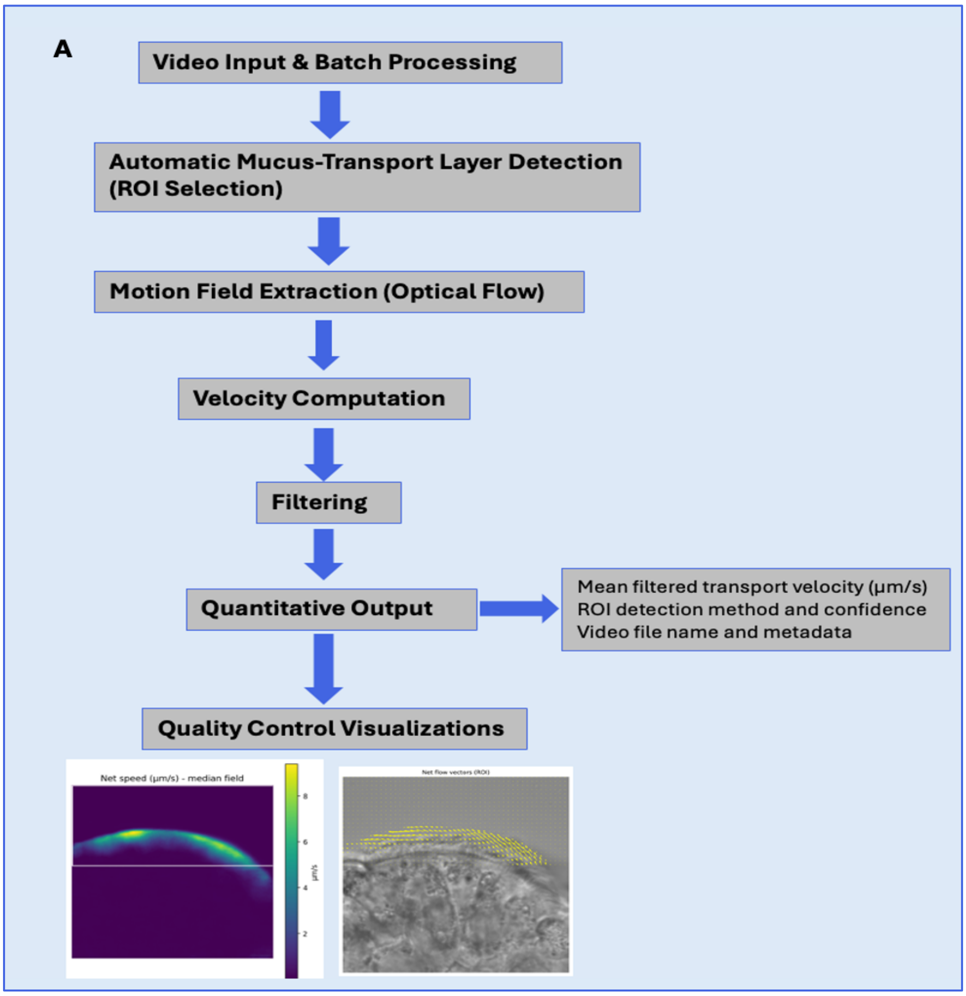


**Supplementary Figure 2 - Workflow of the optical flow clearance method.**

##### Ciliation quantification

To quantify the level of ciliation post culture, cells were scraped and dried on slides overnight. All slides were double labelled with acetylated α-tubulin (T6793, Sigma Aldrich, St. Louis, MO; 1:500) to visualize cilia and with TP63 (Thermo Fisher Scientific, UK; 1:100) to visualize basal cells. Liquid mountant containing DAPI (ProLong Gold P36931, Invitrogen Life Technologies, UK) was used to preserve the slides and to visualize the cell nucleus. Slides were scanned under an inverted confocal laser microscope Leica SP5 (Leica Microsystems Ltd., Milton Keynes, UK) using a x40 objective. For each slide, 5 top views were randomly selected and analysed in Fiji software 2.9.0. The mean fluorescence intensity of acetylated α-tubulin (expressed per RFU: Relative Fluorescent Units) compared to TP63 was automatically obtained for each epithelial strip and mean values were calculated.

##### Whole exome sequencing

Genomic DNA extracted from peripheral blood lymphocyte samples was subject to whole exome sequencing analysis using Agilent enzymatic fragmentation and SureSelect XT HS2 for library preparation with barcoding and pre-capture pooling. Exome libraries were paired-end 150-bp sequenced on an Illumina NovaSeq 6000 system. The alignment of FASTQ files to the human reference genome, variant calling and annotation, followed by variant prioritization and filtration were performed as described previously (PMID: 37077557).
